# Single-Molecule Detection of SARS-CoV-2 by Plasmonic Sensing of Isothermally Amplified Nucleic Acids

**DOI:** 10.1101/2021.10.05.21264561

**Authors:** Haihang Ye, Chance Nowak, Yaning Liu, Yi Li, Tingting Zhang, Leonidas Bleris, Zhenpeng Qin

## Abstract

Single-molecule detection of pathogens such as SARS-CoV-2 is key to combat infectious diseases outbreak and pandemic. Currently colorimetric sensing with loop-mediated isothermal amplification (LAMP) provides simple readouts but suffers from intrinsic non-template amplification. Herein, we report that plasmonic sensing of LAMP amplicons via DNA hybridization allows highly specific and single-molecule detection of SARS-CoV-2 RNA. Our work has two important advances. First, we develop gold and silver alloy (Au-Ag) nanoshells as plasmonic sensors that have 4-times stronger extinction in the visible wavelengths and give 20-times lower detection limit for oligonucleotides than Au nanoparticles. Second, we demonstrate that the diagnostic method allows cutting the complex LAMP amplicons into short repeats that are amendable for hybridization with oligonucleotide-functionalized nanoshells. This additional sequence identification eliminates the contamination from non-template amplification. The detection method is a simple and single-molecule diagnostic platform for virus testing at its early representation.

**Table of Content:** 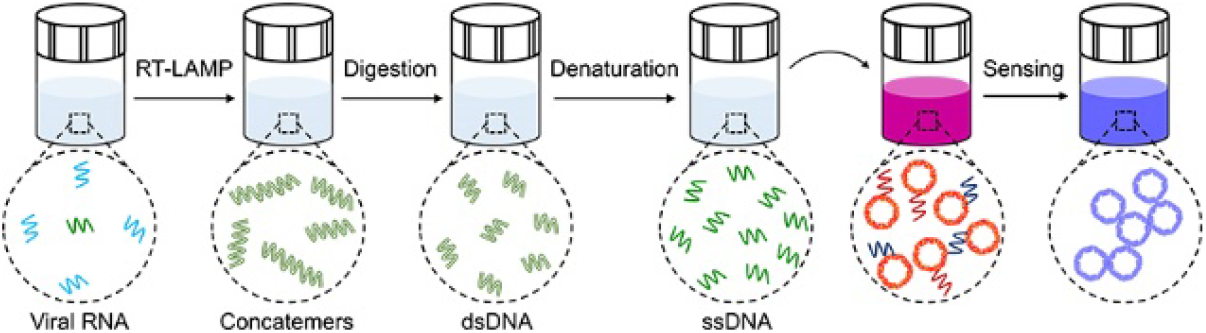

## Introduction

One of the ultimate goals in biological sensing is to detect analytes at single-molecule level.^[1-3]^ For nucleic acid tests, polymerase chain reaction (PCR) has been the gold standard for single-molecule diagnosis.^[3, 4]^ However, its time-consuming protocols and the needs for laboratory infrastructure largely preclude it from point-of-care (POC) testing.^[5, 6]^ Isothermal amplification methods, such as loop-mediated isothermal amplification (LAMP), have emerged as an alternative to PCR and allow POC testing without the need for thermal cycling.^[7]^ Although simple, LAMP is susceptible to non-template amplification and its simple readout (e.g., based on pH change) cannot distinguish template versus non-templated amplification, thus susceptible to false-positive results.^[8-11]^ Previous efforts in implementing gold nanoparticles (AuNPs)-based plasmonic sensing on LAMP products have similar features.^[12-14]^ Specifically, AuNPs have weak optical response and lead to little sensitivity enhancement, while their non-specific sensing of ionic strength change or conjugated primers limits the detection specificity. It is therefore important to develop techniques that can directly identify the amplified sequences via simplified detection scheme, in order to provide diagnostic platforms with simple readouts, high specificity, and single-molecule detection sensitivity.

In this study, we report a robust method for nucleic acid detection, based on plasmonic sensing of LAMP amplicons via DNA hybridization, termed as plasmonic LAMP. Using SARS-CoV-2 RNA as a model, we demonstrate that the plasmonic LAMP method achieved single-molecule detection via colorimetric analysis, which is a desirable diagnostic toolkit to reduce the severity of COVID-19 pandemic.^[9, 15, 16]^ Plasmonic LAMP has two distinctive features. First, we developed Au-silver (Ag) nanoshells as sensitive plasmonic labels. The Au-Ag shells have 4-times stronger plasmonic extinction in the visible wavelengths and provide 20-times sensitivity enhancement in plasmonic sensing of oligonucleotides over AuNPs. Second, we introduced restrict enzyme digestion and heat denaturation in order to cut the concatemer-like LAMP amplicons into short repeats that are amendable for subsequent hybridization with plasmonic sensors (**Scheme 1**). In contrast, previous studies relying on direct sensing of complex LAMP amplicons have met with limited success, due to the lack of hybridization mechanism.^[7, 17, 18]^ The dual sequence identification of plasmonic LAMP, enabled by the primers and plasmonic sensors, eliminates the contamination from non-template amplification and thus improves the detection specificity and sensitivity. Our study is an important step toward advancing simple and ultrasensitive diagnosis.

## Results and Discussion

### Controllable Synthesis and Characterization of Au-Ag Nanoshells

We started with the synthesis of Au-Ag nanoshells through titrating AgNPs with HAuCl_4_. In a standard synthesis, an aqueous solution of HAuCl_4_ is injected at a speed of 6 mL/h into a matrix solution containing AgNPs with an average size of 32 nm as the sacrificial templates (**Figure S1**) and 2 mM sodium citrate (Na_3_CA) as a reductant and a colloidal stabilizer (see **Methods** for details). In this case, AuCl_2_^−^ is first generated from AuCl_4_^−^via Na_3_CA reduction and directs the galvanic replacement reaction with favored high standard reduction potential.^[19]^ **Figure 1A** illustrates the growth of Au-Ag shells, where a pitting process can be observed in the beginning of reaction, followed by the cavity evolution and simultaneous shell formation as more HAuCl_4_ are injected. **Figure 1B-D** show the transmission electron microscopy (TEM) images of products taken from the reaction, confirming the morphological transformation. In contrast, the absence of Na_3_CA leads to the formation of thin and porous cages at a HAuCl_4_ load of 3.3 Ml (**Figure S2**, in equilibrium to 10 mL AuCl_2_^−^). The different products can be attributed to the reaction stoichiometry, where one Ag atom is replaced by one Au atom in the case of AuCl_2_^−^, instead of three Ag atoms for AuCl_4_^−^. Also, the dissolution of the Ag template requires a longer time in the case of AuCl_2_^−^ since less openings are preserved on the surface.^[19]^ The slow pitting process increased shell thickness and thus limits the red shift of the LSPR peak as compared to that of cages (more in next section). The energy-dispersive X-ray (EDX) mapping of an individual Au-Ag shell (**Figure 1E**) shows that the Au and Ag elements are distributed across the entire particle, confirming the alloyed structure. **Figure S3** shows an atomic-resolution high-angle annular dark-field scanning TEM (HAADF-STEM) image of a single particle, where the lattice spacing (1.43 Å) can be attributed to the interplane distance of the (110) planes of the face-centered cubic Au and Ag. The alloy structure is expected to improve the corrosion resistance of Au-Ag nanoshells.^[20]^

**Figure 1.**
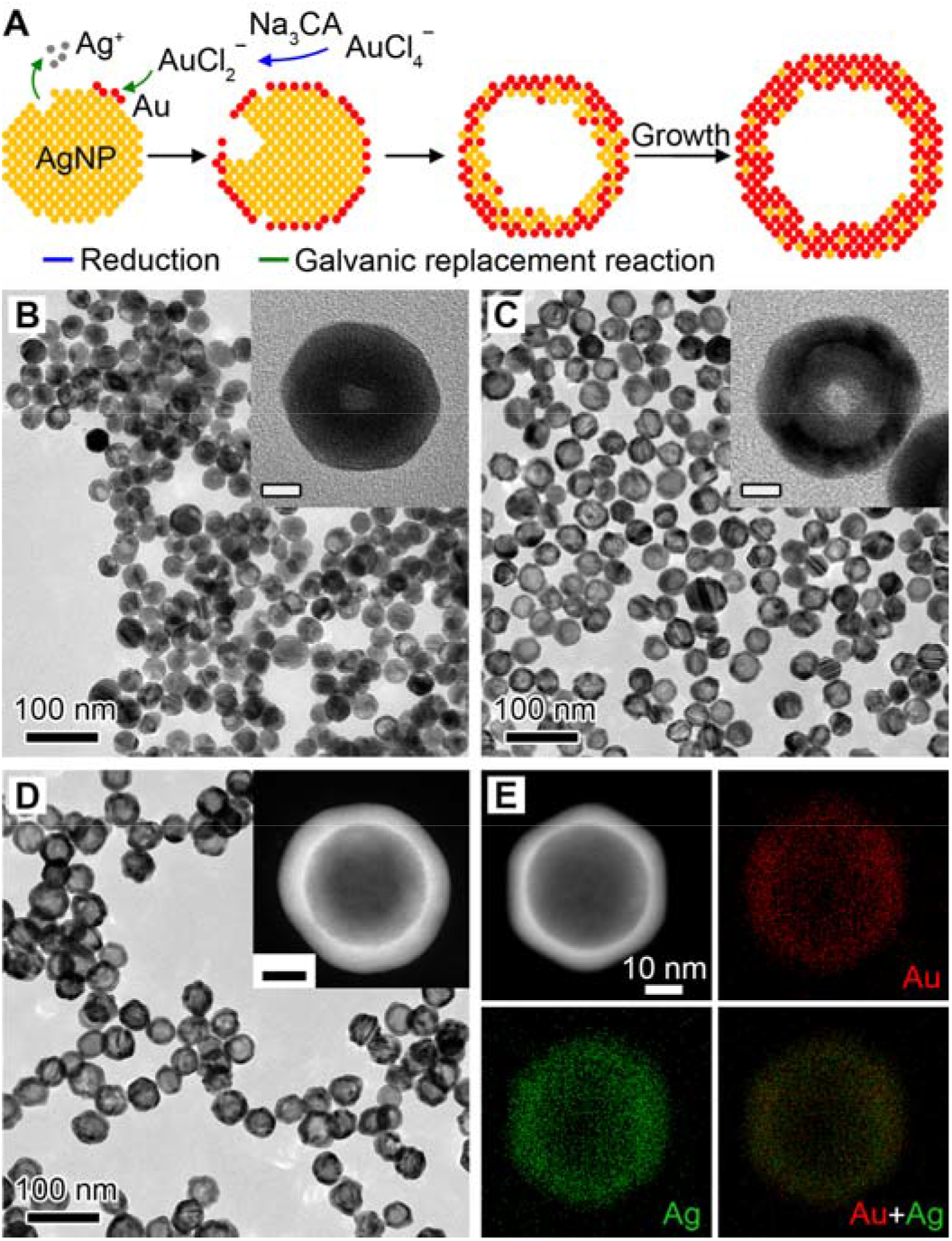
Synthesis and characterization of Au-Ag nanoshells. **A** Schematic illustration of the Au-Ag shells growth based on galvanic replacement reaction in the presence of Na_3_CA. **B-D** TEM images of aliquots taken from the reaction when 1.0 mL (B), 2.5 mL (C), and 10.0 mL (D) HAuCl_4_ was injected. Insets show the magnified TEM images (B, C) and HAADF-STEM image (D) of individual particles. Scale bars in insets are 10 nm. **E** EDX mapping image of an individual particle shown in (D).

We then evaluated the role of Na_3_CA in the shell growth by increasing its concentration from 0 to 20 mM while keeping other conditions the same. Due to the reducing ability endowed by Na_3_CA, we assume that the different reaction species of Au precursor (i.e., AuCl_4_^−^ and AuCl_2_^−^) would influence the reaction stoichiometry (**Figure S4A**). Specifically, without Na_3_CA, the reaction proceeds between AuCl_4_^−^ and AgNPs and results in porous cages. At low concentrations, Na_3_CA turns a small portion of AuCl_4_^−^ into AuCl_2_^−^ and leads to the mixture of cages and shells. In contrast, above a critical concentration (1∼2 mM), Na_3_CA converts the majority of AuCl_4_^−^ into AuCl_2_^−^ and yields shells as the final product. As expected, the synthesized particles (**Figure S4B-G**) have a cage-like morphology below 1 mM Na_3_CA and shell-like structure above 2 mM, confirming the growth mechanism. We also found that the reaction kinetics affect the shell growth (**Figure S5A**). Replacing the Na_3_CA with stronger reducing agent like *L*-ascorbic acid leads to a mixture of small NPs and shells (**Figure S5B**). This is because the *L*-ascorbic acid can also reduce AuCl_4_^−^ into atoms for the self-nucleation growth of NPs. On the other hand, boosting the loading rate of HAuCl_4_ by increasing its concentration and injection speed led to the formation of branched Au-Ag shells (**Figure S5C**). When either the HAuCl_4_ concentration or injection speed increases, Au-Ag shells with a few holes on the surface can be observed (**Figure S5D, E**), suggesting a small portion of AuCl_4_^−^ participated in the reaction. In all cases, the increased deposition rate of Au atoms is key to the preferential overgrowth of branched or porous shells, given that the diffusion rate remains constant during the growth.^[21]^

### LSPR Properties of Au-Ag Nanoshells

Next, we investigated the LSPR properties of the Au-Ag shells since they are critical for plasmonic sensing. **Figure S6** shows the photographs of aliquots taken from the reaction in the presence and absence of Na_3_CA. The suspension of Au-Ag shells remains red-purple, while the cages change from yellow to blue with lower amount of Au precursor injected. The corresponding extinction spectra clearly reveal a different peak shift between these two structures. The LSPR extinction peak (LSPR λ_max_) of Au-Ag shells shifted from 392 nm to 530 nm (**Figure 2A**), while that of cages shifted to longer near-infrared wavelength (**Figure 2B**). The limited and slow peak shifting of Au-Ag shells (**Figure 2C**) arises from the thick and solid shells, as compared to the cages.^[19, 22]^ Furthermore, the extinction intensity change of the cages and shells differs from each other, with a monotonic decrease and falling-rising trend, respectively (**Figure 2D**). This suggests the Au-Ag shells have higher extinction than the cages. Similar trends were observed for the Au-Ag shells and cages synthesized with different Na_3_CA concentrations (**Figure S4, S7**).

**Figure 2.**
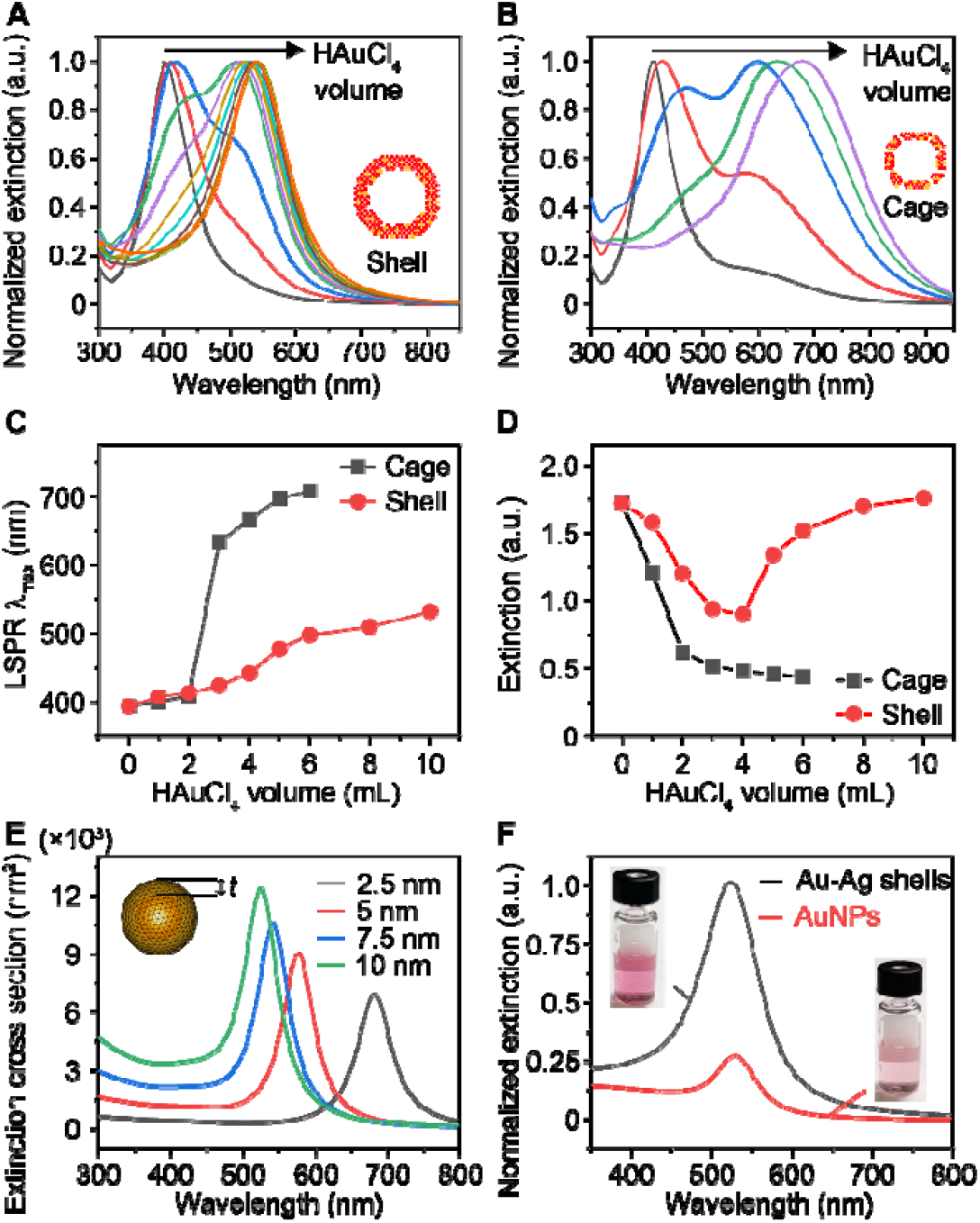
The LSPR properties of Au-Ag nanoshells and nanocages. (A-B) LSPR extinction spectra of aliquots taken from the shell-growth (A) and cage-growth reaction (B) at an injection increment of 1 mL HAuCl_4_. Insets show the models of shell and cage. (C-D) Comparison of LSRP extinction peak λ_max_ (C) and maximal extinction intensity *I*_ext_ (D) change during the growth of shells and cages. (E) Simulation results of the extinction cross-section area of a hollow shell with varied thickness (*t*). Inset shows the model. (F) Normalized LSPR extinction of Au-Ag shells and 50 nm AuNPs at same particle concentration. Insets show the photographs of corresponding suspensions.

To gain further insights into the LSPR properties of the Au-Ag shells, we performed a numerical simulation using the boundary element method.^[23, 24]^ Based on the TEM image of the Au-Ag shell (**Figure 1D**) and the Au: Ag molar ratio of 76%: 24% obtained from inductively coupled plasma-mass spectrometry, we set the geometric model with Au: Ag molar ratio of 80%: 20% (homogeneously distributed), 30 nm interior diameter, and varied thickness (*t*) of wall ranging from 2.5 nm to 10 nm (**Figure 2E** inset). The simulation results suggest that increasing the shell thickness leads to blue shift of LSPR λ_max_ and higher extinction cross-section area (**Figure 2E**). We also found that both the electric field intensity and distribution continuously increase with the shell thickness (**Figure S8**). The enhancement in local field effect of Au-Ag shells is mainly ascribed to the collective effect of surface plasmons of unique shell structure, which condenses the interacting light to the structural center.^[25]^ It should be pointed out that in previous studies, it has been reported that the holes on the surface of cages have a negligible effect on the LSPR λ_max_.^[26]^ This phenomenon is also observed in the shells, as evidenced by the extinction spectra, where the shells with and without holes show close peak positions (**Figure S5F**). Therefore, we did not include this parameter in the simulation. We also compared the LSPR properties of Au-Ag shells and 50 nm AuNPs (**Figure S9**) at the same particle concentration and observed a 4-times higher peak extinction while maintaining similar peak position at visible wavelengths (500-600 nm, **Figure 2F**). Taken together, the Au-Ag nanoshells possess stronger plasmonic extinction at visible range compared with AuNPs and have the potential for be a class of highly sensitive plasmonic sensors.

### Sensitive Colorimetric Detection of Oligonucleotides by Au-Ag Nanoshells

We further examined the Au-Ag nanoshells as labels for oligonucleotide detection. First, a sequence associated with the SARS-CoV-2 N gene (nucleocapsid phosphoprotein gene) was chosen as the target and two complementary sequences were subsequently designed as probes (**Table S1**). Serial dilutions of the target oligonucleotides were mixed with a pair of shells-based sensors in a hybridization buffer and subjected to naked-eye observation or UV-Vis measurements (**Figure 3A** and **S10A**). **Figure S10B** shows the digital photo taken from the completed assay solutions, where a color change can be observed at a target concentration of 250 pM against the blank sample (0 pM) as a reference. For quantitative analysis, the corresponding LSPR extinction spectra were recorded and normalized at 537 nm (**Figure 3B**). A calibration curve was obtained by plotting the extinction intensity (*I*_ext_) ratio between λ_max_ = 675 nm and 537 nm against target concentration (**Figure 3C**). A good linear relationship (*R*^2^ = 0.99) was observed in 1-100 pM (inset of **Figure 3C**). The limit of detection (*LOD*) was determined to be 3.6 pM, by calculating the concentration corresponding to a signal that is 3 times standard deviation above the zero calibrator.^[27]^ Notably, this LOD is approximately 20 times lower than that of using 50 nm AuNPs as probes under same conditions (**Figure S11**). Because the Au-Ag shells and AuNPs have similar size and shape, the improved LSPR property including the 4-fold higher extinction intensity of the shells is likely the key to the sensitivity enhancement. It is worth noting that a ratiometric quantification method is adopted in the present work, which also contributes to the improved analytical performance.

**Figure 3.**
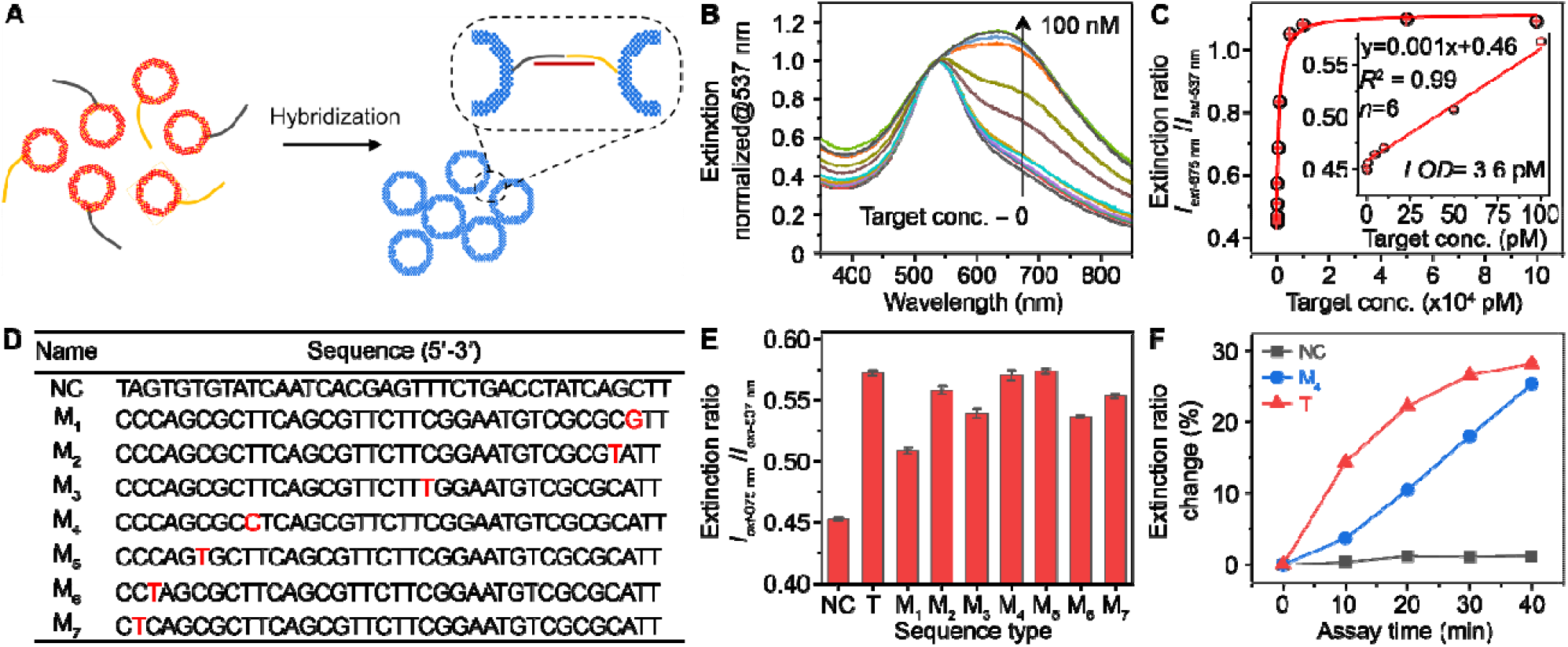
Detection performance of Au-Ag-shells-based plasmonic coupling assay for oligonucleotides. (A) Schematic illustration of plasmonic coupling of Au-Ag shells via DNA hybridization. (B) Normalized LSPR extinction spectrum of the completed assay solutions at varied target concentrations. (C) Corresponding calibration curve generated by plotting the extinction intensity ratio against target concentration. A logistic fitting is applied. Inset shows inear region of the calibration curve. (D) A table of sequence information for non-complementary oligonucleotide (NC) and single base-pair mismatched oligonucleotides (M_1_-M_7_). NC sequence is selected from a malarial sequence. Red letter in mismatched sequences highlights where the single base-pair mismatch occurs. (E) Optical responses of the shells-based probes to NC, target (T), and M_n_ (n=1-7) oligonucleotides. (F) Spectral monitoring of the hybridization kinetics for different types of oligonucleotides. All sequences had concentration of 100 pM. The extinction ratio (*R*_ext_) change is calculated by (*R*_ext-t min_ - *R*_ext-0 min_) / *R*_ext-0 min_ × 100%, where *R*_ext-t min_ = *I*_ext 675 nm-t min_ / *I*_ext 537 nm-t min_.

We further assessed the detection selectivity by testing the probes with a variety of oligonucleotide sequences, including a non-complementary sequence (NC), a perfectly matched target sequence (T), and a group of single-base-pair (SBP) mismatched sequences (M_n,_ n=1-7, see details in **Figure 3D**). The M_n_ sequences are taken from the SARS-CoV-2 Browser and have been confirmed to be polymorphism variants isolated in the US.^[28]^ Therefore, an assay with SBP-mismatched tolerance is of paramount importance since false-negative results can cause delayed treatment and increased risk of viral transmission. We tested sequences with these 7 different mutations and found that all showed positive detection above the negative control (**Figure 3E**). This result suggests a good tolerance for SBP mismatch. The slight difference among M_n_ sequences may result from the mismatch locations and the direct hybridization mechanism.^[29]^ Careful examination shows that monitoring the hybridization kinetics at room temperature can distinguish the SBP mismatch from the positive control sequence without mutations (**Figure 3F**) and provides a potential way to further detect SBP mismatch.

### Single-Molecule RNA Detection by Plasmonic LAMP

Lastly, we developed a diagnostic protocol to integrate reverse transcription loop-mediated isothermal amplification (RT-LAMP) with plasmonic Au-Ag nanoshells for single-molecule detection of infectious pathogens such as SARS-CoV-2 (**Scheme 1**). We first designed primers targeting six sites flanking a conserved region in the N gene of SARS-CoV-2 (**Figure 4A**, sequence information in **Table S2**). With RT-LAMP, the viral RNA was reverse transcribed into complementary DNA and subsequently amplified via self-priming loop structures and a high strand-displacing DNA polymerase that generates large cauliflower-like concatemers.^[7]^ The RT-LAMP products were verified by agarose gel electrophoresis revealing multiple bands in a ladder-like pattern indicative of successful LAMP amplification, whereas a sample without RNA input lacked any such bands (red box of **Figure 4B**). The concatemers were then subjected to restriction endonuclease digestion by HincII and EaeI, which recognize and cleave GTY/RAC and Y/GGCCR sites, respectively. These sites are conserved throughout the long concatemers such that their cleavage by digestion collapses the concatemer bands and produces significantly shorter sequences more suitable for detection. **Figure 4B** (blue box) reveals clean bands located below 200 base-pair (BP) length, confirming the uniform digestion. Next, the reaction product was denatured into single-strand DNA (ssDNA) by heating to 95°C and cooling on ice, followed by mixing with a set of shells-based probes and incubated at 62 °C for 10 min or room temperature for 30 min. Significantly, a color change can be observed at RNA input ≥10 copies/µL (**Figure 4C**). **Figure 4D** shows corresponding LSPR extinction spectra normalized at 537 nm. A linear relationship (R^2^ = 0.99) was observed covering 3 logs from 1 to 1,000 copies/µL (**Figure 4E**). The LOD was determined to be 1 copy/µL and confirmed the capability of plasmonic LAMP approach for the single-molecule RNA detection. When used respiratory syncytial virus (RSV) RNA as initial target, it led to insignificant spectral change as compared to the reference sample, indicating the high specificity of this assay. The protocol developed here only requires easily accessible heat blocks and is suitable for POC testing.

**Figure 4.**
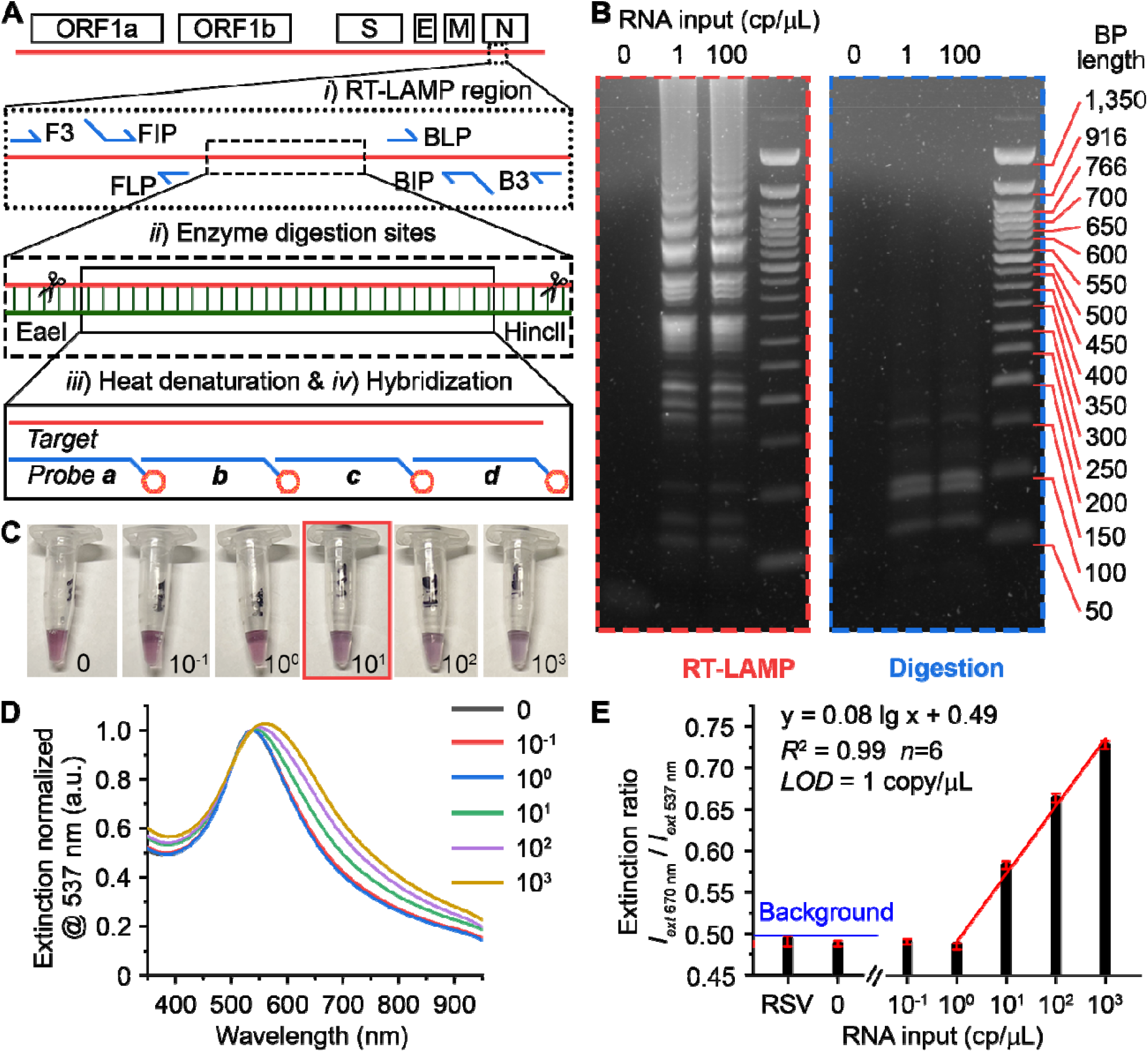
Plasmonic LAMP for single-molecule detection of SARS-CoV-2 RNA. (A) Schematic illustration of the SARS-CoV-2 genome architecture and reaction proceeding. Target region, primers, cutting sites, and probe sequences are highlighted. (B) Gel images showing the representative products after RT-LAMP and enzyme digestion at different RNA input. (C) Photographs taken from the completed assay at varied RNA input (copies/µL). The red box highlights the visual detection limit. (D) Corresponding LSPR extinction spectra (normalized at 537 nm) of the detection results shown in (C). (E) Linear region of the calibration curve shown in (D). Error bars indicate the standard deviations of six parallel measurements. RSV is respiratory syncytial virus, and its RNA was extracted and cleaned before use.

It should be emphasized that the enzyme digestion and heat denaturation (steps ***ii*** *and* ***iii*, Figure 4A**) involved in the plasmonic LAMP are necessary. Without step ***iii***, directly detecting the RT-LAMP products via oligonucleotide sensors results in no color change since RT-LAMP amplicons are double-strand DNA (dsDNA) with no available binding sites (sample **c, d, Figure S12**). Without step ***ii***, the produced ssDNA after heating could easily re-anneal back as dsDNA in the hybridization buffer due to the extensive stem-loop and cauliflower-like structure of the RT-LAMP products.^[30]^ Likewise, this would prevent the binding of the NP probes and does not lead to color change (sample **e, Figure S12**). In contrast, with step ***ii*** and ***iii***, the dsDNA can be cut into uniform and short structures, which are amendable for subsequent hybridization detection with short oligonucleotides as probes, leading to highly specific and sensitive detection.^[7]^ In contrast, previous reports rely on indirect sensing, where the ionic strength change in a salt solution or molecules labelled on primers essentially induce the aggregation of AuNPs-based sensors.^[12-14, 31-33]^ The non-template amplification could still lead to false-positive results.

To demonstrate the advances of plasmonic LAMP over commercially available colorimetric LAMP kit, we compared their detection performance for SARS-CoV-2 RNA. **Figure 5A** shows the photographs of colorimetric LAMP incubating with different SARS-CoV-2 RNA inputs and RSV RNA as a negative control. The reaction was conducted for up to 45 min at 65°C. It can be seen that it only shows color change for SARS-CoV-2 RNA molecules of 50 copies/µL after 30 min incubation, while no color change can be observed for a short reaction time (e.g., 15 min). In contrast, at time point of 45 min, all samples turn into yellow color, resulting in undistinguished positive and negative controls. This is indeed caused by the non-template amplification products and thus compromise the detection sensitivity.^[9, 10]^ We then performed plasmonic LAMP for the samples processed with 15 min and 45 min amplification. Spectral analysis of the completed assay solutions suggests that plasmonic LAMP pushes down the detection limit to 50 copies/µL at 15 min and discriminates signals for 5 copies/µL against control sample at 45 min (**Figure 5B**). Evidently, plasmonic sensing enhances the detection sensitivity and specificity of LAMP and in turn makes the plasmonic LAMP a reliable single-molecule diagnostic approach for nucleic acid detection.

**Figure 5.**
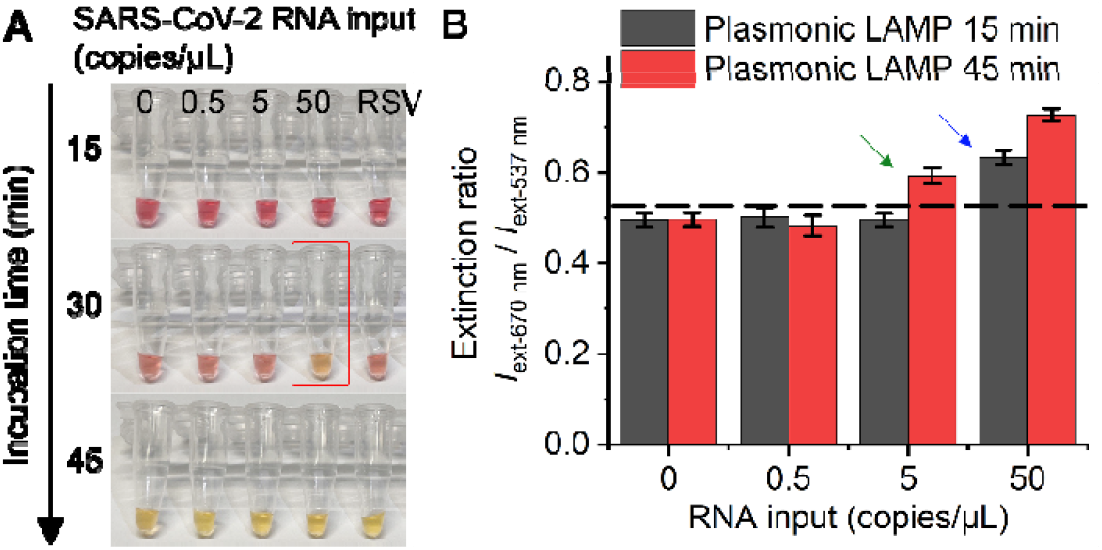
Comparison between plasmonic LAMP and a commercially available colorimetric LAMP kit for SARS-CoV-2 RNA detection. (A) Photograph of colorimetric LAMP reaction with different RNA inputs and incubation time at 65 °C. Red box highlights the detectable sample. RSV RNA (50 copies/µL) was used as negative control. (B) Detection results of plasmonic LAMP at different incubation times. At indicated times, LAMP products were processed for plasmonic sensing using Au-Ag-shells-based probes. Dashed line marks the background (3 times standard deviation above mean). Blue and green arrows indicate the detection limit for 15 min and 45 min, respectively.

To demonstrate the potential clinical use, we applied the integrated plasmonic LAMP approach to detect nasal swab samples that were spiked with SARS-COV-2 RNA. RNA of six different concentrations in the range of 0-100 copies/µL were spiked in healthy nasal swab samples and tested. The analytical performance shows sensitive detection (**Table 1**) and suggests that the Au-Ag-shells-based sensors work well in a complex sample matrix and have potential uses in clinical setting. Taken together, the established diagnostic approach should be readily adopted as a nucleic-acid detection platform simply by changing the set of primers and probing sequences.

**Table 1.** Analytical Performance of Plasmonic LAMP Detection of SARS-CoV-2 RNA-Spiked Nasopharyngeal Swab Samples.

| Viral copies in sample<br>(copies/μL) | Plasmonic LAMP |
| --- | --- |
| 100 | 6/6<sup>[a]</sup> |
| 50 | 6/6 |
| 10 | 6/6 |
| 5 | 4/6 |
| 1 | 1/6 |
| 0 | 0/6 |

[a]: Total positive samples versus total testing sample.

## Conclusions

In summary, we have developed the plasmonic LAMP approach for the single-molecule detection of SARS-CoV-2 RNA. With enzyme digestion and heat denaturation, plasmonic LAMP allows identifying amplicons via DNA hybridization utilizing Au-Ag nanoshells as sensors, yielding detection limit at 1 RNA copy per microliter. The Au-Ag shells can be prepared from AgNP templates through titration with Au^+^ ions and have thick and hole-free surface. They show LSPR peak around 530 nm and high peak extinction, offering substantially enhanced detection sensitivity (20-times) compared to that of conventional AuNPs-based assay. Plasmonic LAMP has improved detection performance and only requires easily accessible heat blocks. Our work provides a diagnostic toolkit with simple readouts and ultralow detection limit that has potential for clinical applications.

## Data Availability

The data are available on request from the corresponding authors.

## Acknowledgements

This work was supported in part by National Institutes of Health (NIH) grant R01AI151374, and U.S. Department of Defense (DOD) grant PR192581, NSF grant 1361355 and Cecil H. and Ida Green Endowment. We would like to specifically thank Dr. Qingxiao Wang and Prof. Moon J. Kim from the University of Texas at Dallas for their general supports in taking the high-resolution electron microscopy images for us.

---

**Scheme 1.**
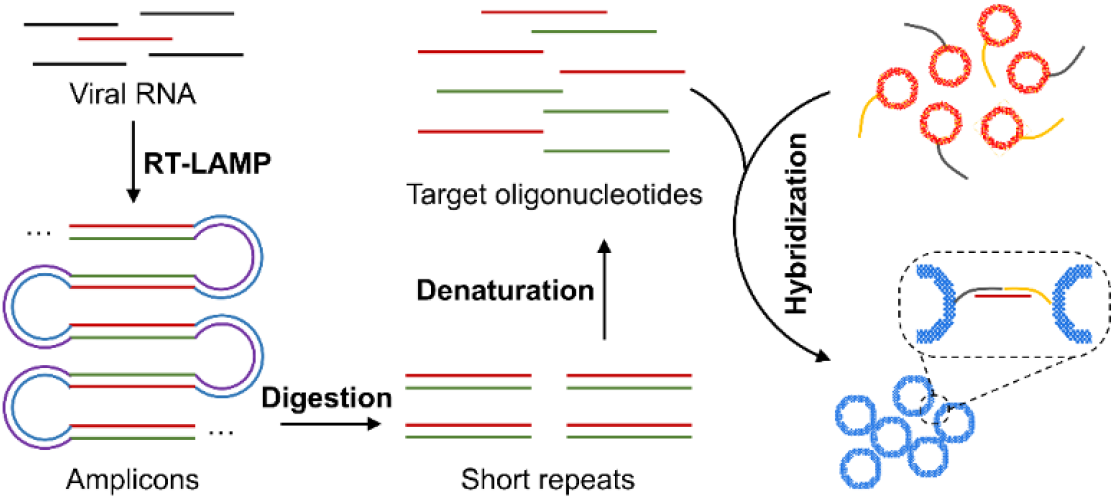
Schematic illustration of the major steps for the proposed plasmonic LAMP single-molecule RNA detection. The viral RNA is first reverse transcribed and amplified into amplicons that are subjected to restriction enzymes digestion, forming short repeats that can be denatured into oligonucleotides for subsequent DNA hybridization linked with plasmonic sensors.

