## Supplemental Materials for "Single-Molecule Detection of SARS-CoV-2 by Plasmonic Sensing of Isothermally Amplified Nucleic Acids"

Experimental Procedures

*Chemicals and Materials.* Gold (III) chloride trihydrate (HAuCl_4_·3H_2_O), silver nitrate (AgNO_3_), sodium citrate dihydrate (Na_3_CAꞏ2H_2_O), sodium borohydride (NaBH_4_). *L*-ascorbic acid (AA), gold and silver standards, tuning solution for ICP-MS tests, nuclease-free water were ordered from FisherScientific Inc. HPLC purified probe sequences for oligo and RNA detection and oligo target sequences including the non-complementary sequence and single base-pair mismatch sequences were ordered from BioBasic Inc. (Markham, ON, Canada). Primers for loop-mediate isothermal amplification were ordered from Sigma-Aldrich (St. Louis, MO, USA). Sherlock CRISPR SARS-CoV-2 were ordered from Integrated DNA Technologies, Inc. Synthetic RNA positive controls were Twist Bioscience SARS-CoV-2 RNA Control 2 ordered from Fisher Scientific Inc. Restriction enzymes (EaeI and HincII) and WarmStart® Colorimetric LAMP 2X Master Mix (DNA & RNA) were ordered from New England Biolabs Inc. Hydroquinone, formamide, dextran sulfate, and sodium chloride (NaCl) were purchased from Sigma-Aldrich (St. Louis, MO, USA). The procured chemicals were used as received. All aqueous solutions were prepared using deionized (DI) water with a resistivity of 18.0 MΩ·cm. All glassware for the synthesis used in the nanoparticle synthesis was cleaned using aqua regia (3:1 ratio of hydrochloric acid and nitric acid).

*Preparation of 4 nm Ag nanoparticles as seeds.* In a typical synthesis, ∼4 nm AgNPs could be prepared by reducing aqueous AgNO_3_ solution with NaBH_4_ according to a previous report.^[1]^ Briefly, 2 mL of 1% (w/v) Na_3_CA solution and 6 mL of DI water were added to a 20 mL vial and preheated at 70 °C in an oil bath under magnetic stirring for 15 min, followed by sequentially adding 0.17 mL of 1% (w/v) AgNO_3_ solution and 0.2 mL of 0.1% (w/v) NaBH_4_ solution. The reaction was kept at 70 °C under vigorous stirring for 1 h. After cooling down to room temperature, the ∼4 nm AgNPs were diluted to 10 mL using DI water and stored in dark for further use.

*Preparation of 32 nm Ag nanoparticles as templates.* 32 nm AgNPs were prepared based on a seeded-growth method.^[2]^ In brief, 5 mL of the prepared ∼4 nm AgNPs as seeds, 1 mL of 1% (w/v) sodium citrate aqueous solution, 1 mL of 1% (w/v) AA aqueous solution, and 35 mL of DI water were mixted in a 100 mL flask and preheated at 80 °C in an oil bath under magnetic stirring for 15 min. Then, 0.85 mL of 1% (w/v) AgNO_3_ solution was added into the mixture using a pipette. The reaction was kept at 80 °C under vigorous stirring for 1 h. The product was centrifuged and washed with DI water three times, redispersed in 50 mL of DI water, and stored in the dark for further use (0.224 nM in particle concentration).

*Preparation of ~50 nm Au-Ag nanoshells.* ~50 nm AuAg nanoshells were prepared via galvanic replacement reaction. In brief, 3 mL of the prepared ∼32 nm AgNPs as templates, 1 mL of sodium citrate aqueous solution with varied concentration (0-10 mM), and 6 mL of DI water were mixed in a 50 mL flask and preheated at 95 °C in an oil bath under magnetic stirring for 15 min. Then, 0.004% (w/v) HAuCl_4_ solution was injected into the mixture using a syringe pump at a speed of 6 mL/h for 10 mL. After injection, the reaction was kept at 95 °C under vigorous stirring for 10 min. The product was centrifuged and washed with DI water three times, redispersed in 3 mL of DI water, and stored in the dark for further use.

*Preparation of ~50 nm Au nanoparticles.* 15 nm AuNP was first synthesized according to our previous publication and subsequently used as seeds for the larger AuNP synthesis.^[3]^ Briefly, water was heated to boiling under magnetic stirring. Immediately after, 1% (w/v) Na_3_CAꞏ2H_2_O and 0.01% (w/v) HAuCl_4_·3H_2_O were sequentially added. After a prominent color change from purple to red, the solution was heated for additional 30 min before cooling down to room temperature. 50 nm AuNPs were synthesized using hydroquinone as a reducing agent. Specifically, hydroquinone was added to a solution of HAuCl_4_·3H_2_O, Na_3_CAꞏ2H_2_O, and the 15 nm AuNPs (used as seed particles) at room temperature. This solution was stirred continually overnight to allow for AuNPs growth.

*Preparation of nanoparticle-oligonucleotide as probes.* Nanoparticle-oligonucleotides conjugation was performed according to our previous work.^[3]^ The poly A-tail probe oligonucleotides were first resuspended in DI water per vendor suggestions before use. The SH-capped oligonucleotides were conjugated separately to nanoparticles in acidic buffer. Briefly, nanoparticle suspension was mixed with oligonucleotide solution before adding a 50 mM citrate-HCl buffer with pH of 3.0 ± 0.1 in a 1:1 volumetric ratio. After 30 min of incubation at room temperature, the nanoparticle-oligonucleotides conjugates were centrifuged and washed with DI water for three times. The purified products were redispersed in DI water and stored in the 4 ^o^C refrigerator for further use.

*Plasmonic coupling assay of oligonucleotide**.* The assay was performed according to our previous work.^[3]^ In a standard approach, a hybridization buffer (20% formamide, 16% dextran sulfate, and 0.6 M NaCl solution) was mixed with probe A and B solution in a volume ratio of 4:3:3. The freshly prepared working solution was then added to the target samples at different concentrations (volume ratio = 2:1). The solution was then incubated at 62 ^o^C for 10 min or room temperature (22 ^o^C) for 30 min prior to the UV-Vis measurement.

*Single-molecule detection of SARS-CoV-2 RNA.* The protocol was performed per the manufacture’s recommendation. In a standard approach, 8 µL of RNA sample with varied copy number is first mixed with RT-LAMP mixture containing 10 µL of 2x Warm Start RT-LAMP mix and 2 µL of 10x customized primers, followed by incubation at 62 ^o^C for 40 min. 2 µL of the product is loaded into a solution containing 1.25 µL of EaeI, 1.25 µL of HincII, 5 µL of rCutSmart™ buffer, and 40.5 µL of water. The mixed sample is incubated at 37 ^o^C for 30 min and then heated to 95 ^o^C for denaturation. Afterwards, the sample is quickly cooled down on ice and ssDNA is formed. To process the plasmonic coupling assay, 50 uL of working solution containing probe suspensions a-d, and hybridization buffer with volume ratio of 1.5:1.5:1.5:1.5:4 was first prepared. Then 25 uL of the ssDNA was was mixed with the working solution and incubated at 62 ^o^C for 10 min or room temperature (22 ^o^C) for 30 min before the UV-Vis measurement or naked eye discrimination.

*RNA extraction and purification of respiratory syncytial virus (RSV).* RSV RNA as a negative control was extracted using a commercially available kit (Viral RNA Extraction Buffer, VRE100, Millipore-Sigma). Briefly, 10 µL viral stock (e.g., 10^3^ PFU/mL as pre-quantified by plaques assay) was mixed with 5 µL extraction buffer at room temperature for 10 min and subjected to purification. A commercially available kit (Monarch® Kits for RNA Cleanup, New England Biolabs Inc.) was used as per the manufacturer’s protocol. The concentration of RNA was then quantified to be 5.9 ng/µL using NanoDrop 2000 (Thermo Scientific^TM^).

*Gel image analysis.* For gel electrophoresis, a 2% agarose gel was made with Low-EEO Agarose powder (FISHER BP160-500) dissolved in 1X Tris-Borate-EDTA (TBE) solution (FISHER BP1333-1) and stained with 0.5 μg/mL of Ethidium Bromide (FISHER BP1302-10). The gel ran at 150V in a 1X TBE buffer solution for 30 minutes powered by a Bio-Rad PowerPac™ Basic (300V 400mA 75W) power supply.

*Clinical-mock sample tests.* Nasal swab samples were collected from a healthy individual with SARS-CoV-2 negative results. BD universal viral transport medium are gifts from UT Southwestern Medical Center and used as collecting buffer. The medium is first pipetted out carefully and loaded with varied amount of RNA. The samples were freshly prepared before use. To proceed the sample test, sample protocol was used same as for RNA detection as mentioned above. The use of human nasal swab sample was approved by Institutional Review Board (IRB) at University of Texas at Dallas (ID: 20MR0093).

*Boundary Element Method (BEM) simulation.* The plasmonic properties of Au-Ag nanoshells were simulated by means of the BEM, which solves Maxwellʼs equations for the geometry of a metallic nanoparticle only in terms of the boundaries that separate different media described by homogeneous and isotropic dielectric functions. All simulations were conducted via the MNPBEM Matlab toolbox.^[4]^ The dielectric functions of silver and gold alloy were obtained from a modified Drude-Lorentz model developed by Rioux *et* *al*.^[5]^ In the extinction spectra and electromagnetic field simulation, the Au-Ag shell model was built to be immersed in water as dielectric background. The electric field maps were computed from the surface charges where the particles were excited by plane wave at wavelength of 532 nm.

*Characterizations.* The extinction spectra in microtiter plates were read using microplate reader (Synergy 2, BioTek). The TEM images were taken using a JEOL JEM-2010 microscope operated at 120 kV. The pH values of buffer solutions were measured using a pH Meter (Accumet AP71). Agarose gel imaging was performed using a Bio-Rad Molecular Imager® ChemiDoc™ XRS+. A FEI 200 kV Titan Themis scanning TEM was used to acquire the HADDF-STEM images and energy dispersive X-ray (EDX) mapping images. A Perkin-Elmer Sciex Elan 6100 DRC inductively coupled plasma mass spectrometer (ICP-MS) was used to determine the amounts of Ag and Au elements in various nanostructures. Gel image analysis was performed using Bio-Rad Image Lab v5.2.1 software. Digital photographs were taken by iPhone 12 ProMax.

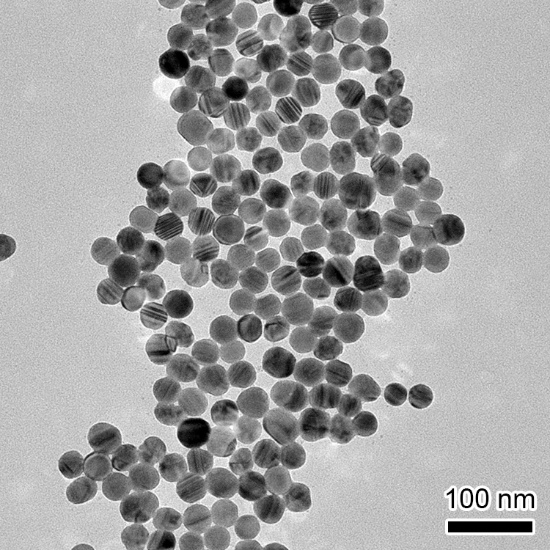

**Figure S1**. Transmission electron microscopy (TEM) image of AgNPs as templates.

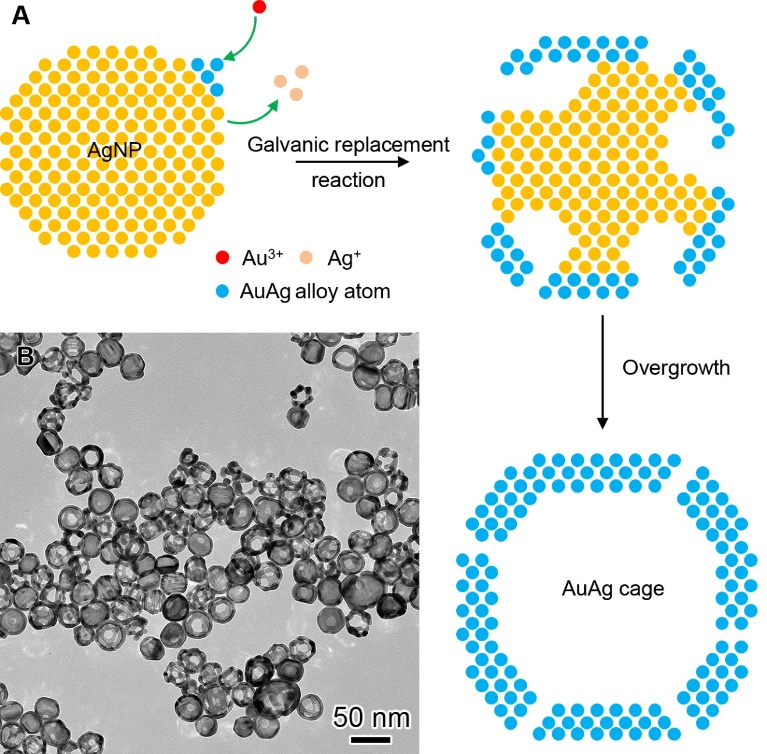

**Figure S2**. **The preparation of Au-Ag nanocages.** (A) Schematic illustration of the cage growth. (B) TEM image of Au-Ag cages obtained at 0 mM Na_3_CA with 3.3 mL HAuCl_4_ injection.

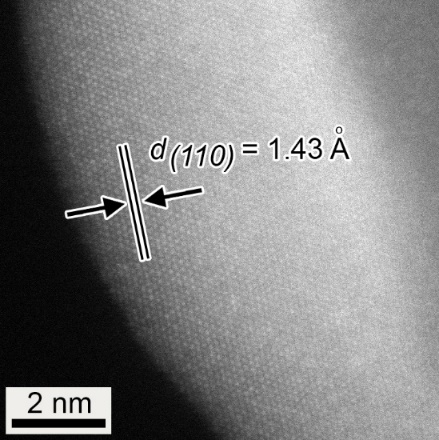

**Figure S3**. **Atomic-resolution high-angle annular dark-field scanning TEM**. The lattice spacing was measured to be 1.43 Å, corresponding to the (110) plane of Au and Ag alloyed structure.

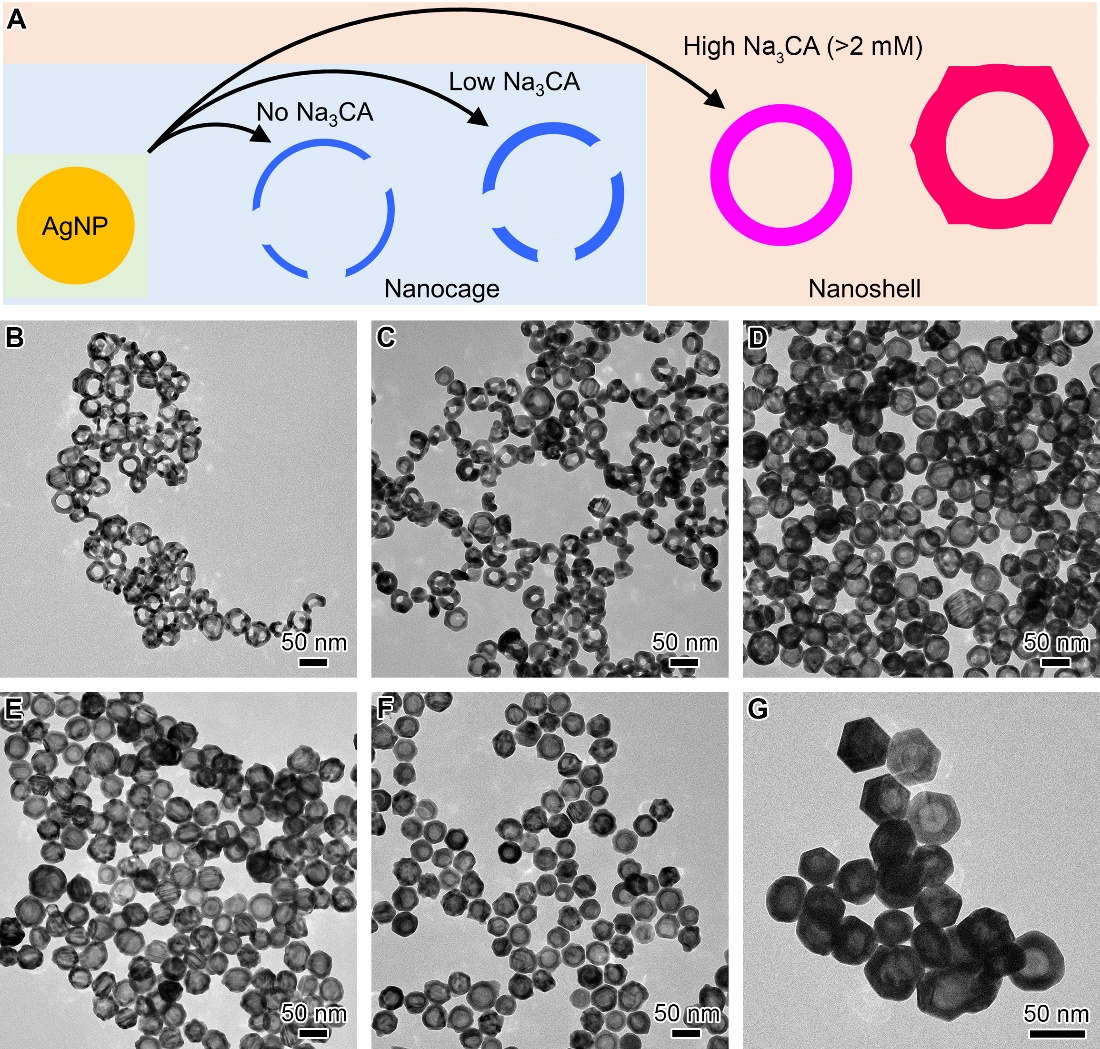

**Figure S4**. **Morphological characterization of hollow Au-Ag nanostructures obtained at varied concentration of Na_3_CA.** (A) Schematic illustration of the growth pathways. (B-G) TEM images of the products obtained in the presence of 0 mM (B), 0.5 mM (C), 1 mM (D), 5 mM (E), 10 mM (F), and 20 mM (G) Na_3_CA. In (B), 6 mL of HAuCl_4_ was injected, while 10 mL HAuCl_4_ was added for the rest cases.

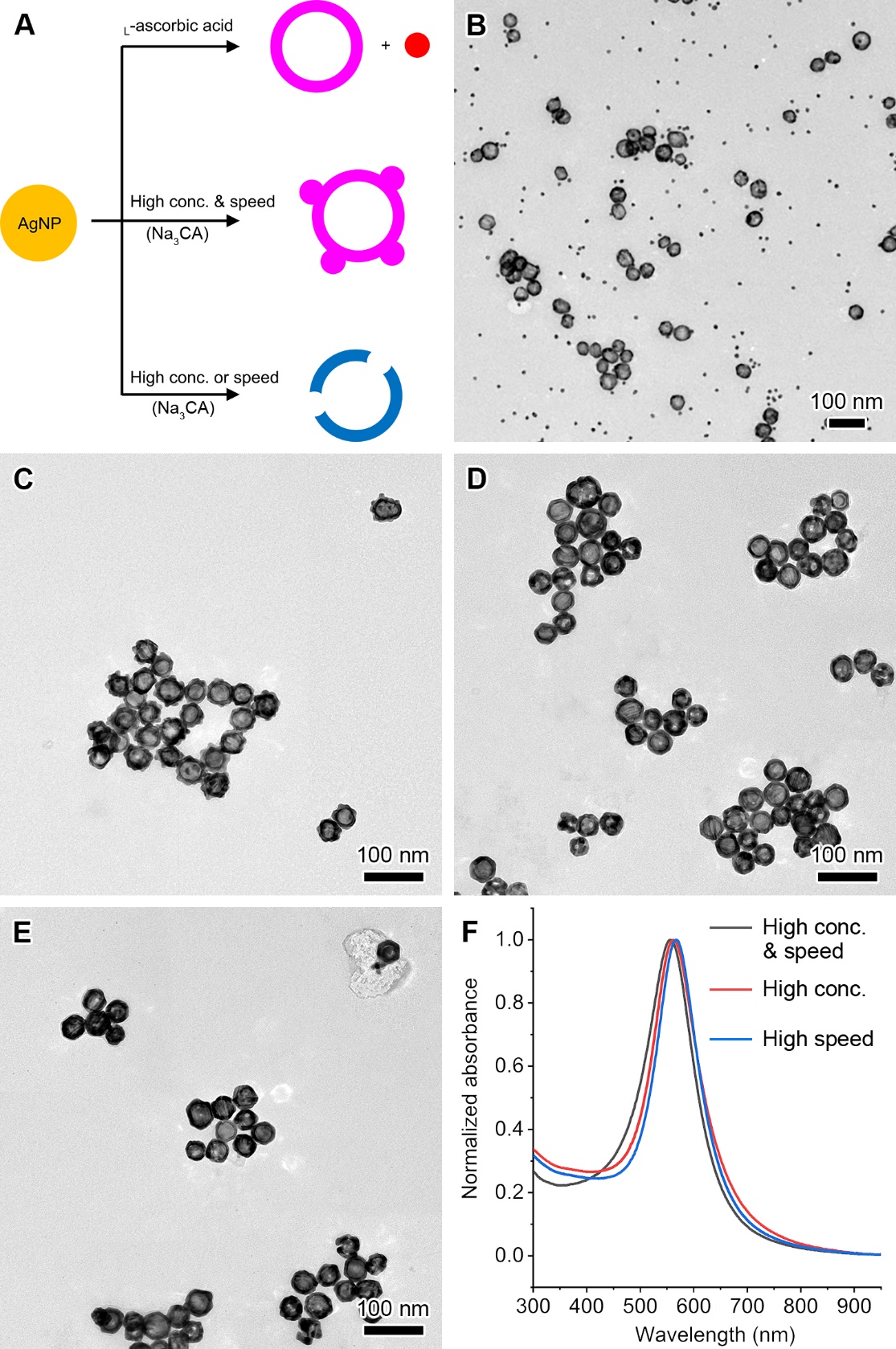

**Figure S5**. **Morphological and optical characterizations of hollow Au-Ag nanoshells obtained at varied conditions.** (A) Schematic illustration of the different growth pathways. (B-E) TEM images of the products obtained, when using 2 mM ascorbic acid as a reducing agent (B), 0.04% (w/v) HAuCl_4_ at 30 mL/h injection speed (C), 0.04% (w/v) HAuCl_4_ at 6 mL/h injection speed (D), and 0.004% (w/v) HAuCl_4_ at 30 mL/h injection speed (E). (F) Normalized extinction spectra showing the corresponding LSPR peaks of the products in (C-E).

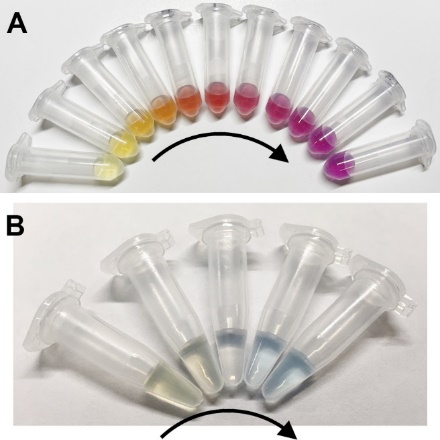

**Figure S6.** Photographs taken from the aliquots during the synthesis of Au-Ag nanoshells (A) and nanocages (B) NCs in the presence and absence of Na_3_CA, respectively. The arrows mark the color change when an increasing amount (per mL) of HAuCl_4_ was injected.

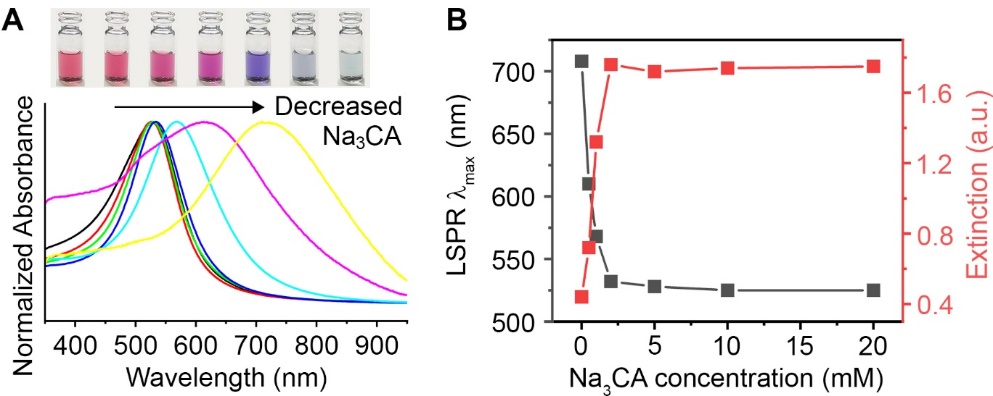

**Figure S7. LSPR properties of hollow Au-Ag nanostructures shown in Figure S4.** (A) Photographs and corresponding extinction spectra of those nanostructures. (B) A plot of the major LSPR peak λ_max_ (LSPR λmax) in (A) against the concentration of Na_3_CA present in the reaction.

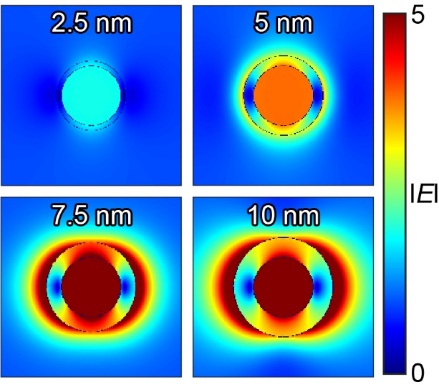

**Figure S8.** Electric field intensity maps (logarithmic scale) of the shells with varied thickness. The map was generated at the major LSPR extinction peak at 532 nm.

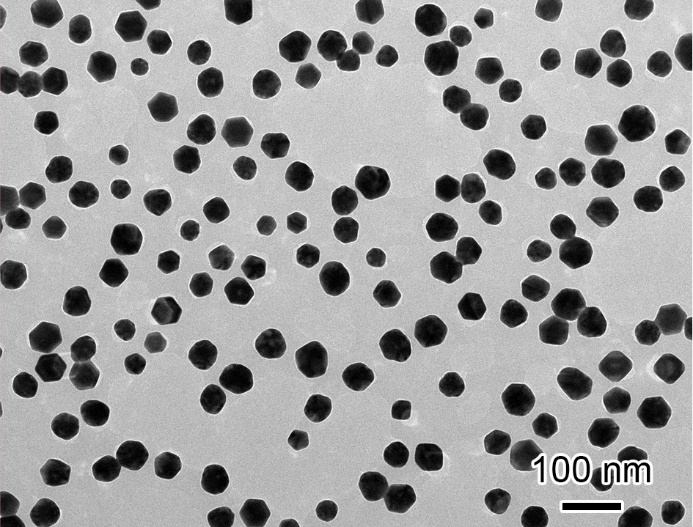

**Figure S9**. TEM image of 50 nm AuNPs.

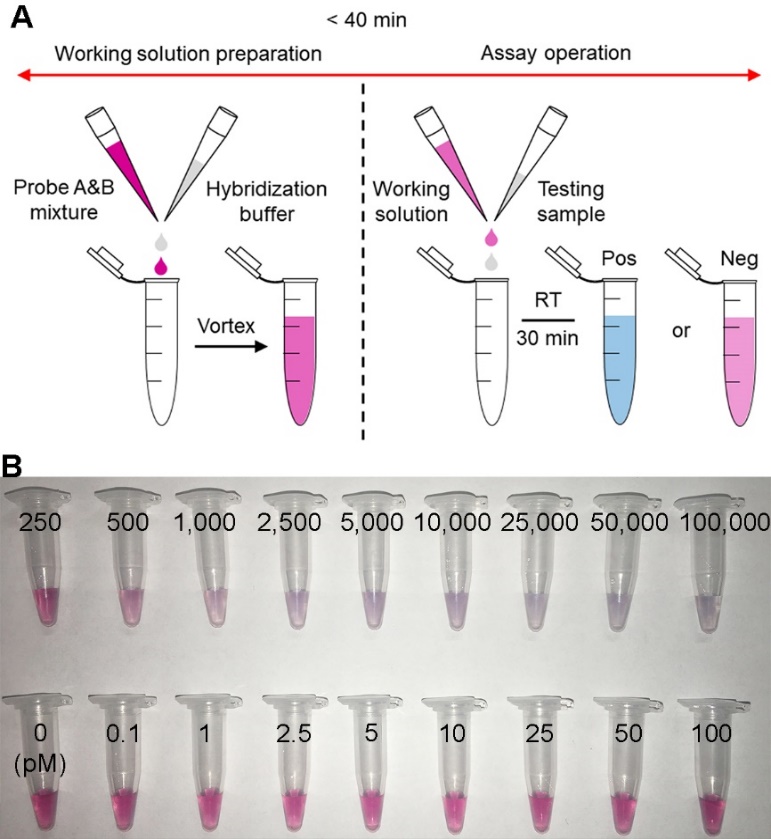

**Figure S10. Plasmonic coupling assay of oligonucleotides using Au-Ag-shells-based sensors. A** Schematic illustration showing the PCA of oligonucleotides using Au-Ag nanoshells as sensors. **B** Representative photographs of the PCA of target standards.

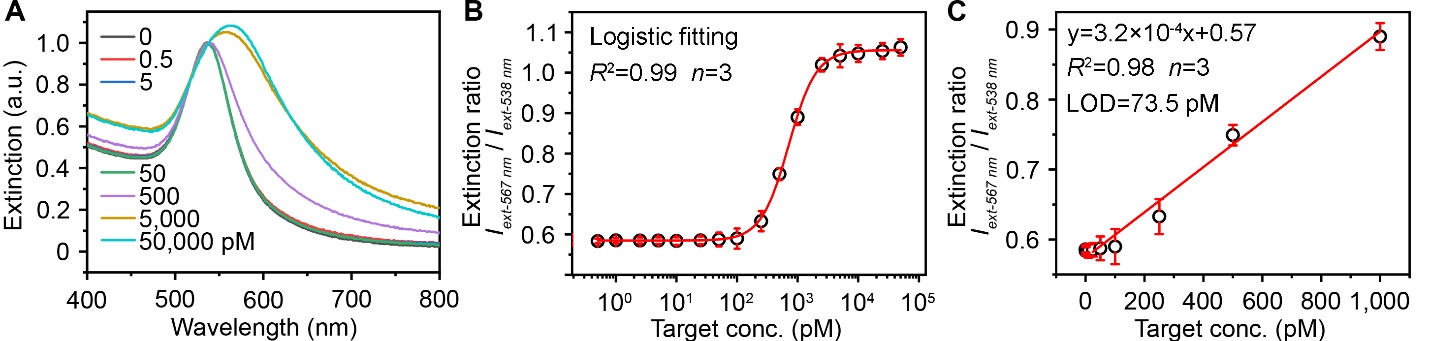

**Figure S11**. **Plasmonic coupling assay of oligonucleotides using** **using 50 nm AuNPs as sensors.** (A) Representative extinction spectra normalized at 538 nm taken from the assay solution with varied target concentrations. (B) Calibration curve generated by plotting the extinction ratio of *I_ext-567 nm_* and *I_ext-538 nm_* against target concentration. A logistic fitting is applied. (C) A linear range region of the calibration curve shown in (C). Error bars indicate the standard deviations (*n* = 3).

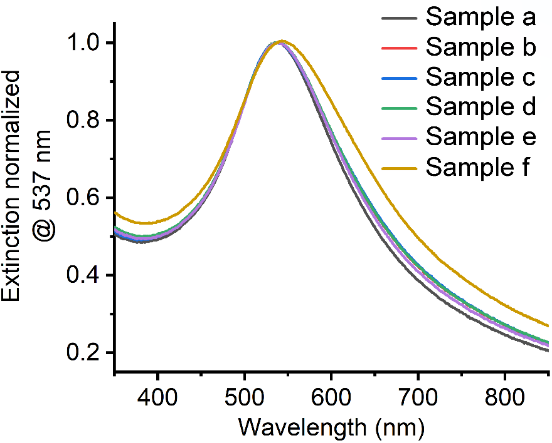

**Figure S12. Plasmonic LAMP performance at varied conditions.** Detailed conditions can be found in below Table.

| **Sample** | **Experimental conditions** | | | | | **Color change** |
| --- | --- | --- | --- | --- | --- | --- |
|  | **RT-LAMP** | **Enzyme digestion** | **Heating denaturation** | **PCA** | **RNA input (cp/µL)** |  |
| **a** | No | No | No | Yes | 0 | ^[a]^No |
| **b** | Yes | No | No | Yes | 0 | No |
| **c** | Yes | No | No | Yes | 10 | No |
| **d** | Yes | Yes | No | Yes | 10 | No |
| **e** | Yes | No | Yes | Yes | 10 | No |
| **f** | Yes | Yes | Yes | Yes | 10 | Yes |

[a]: This sample is used as a reference to indicate the color change.

**Table S1**. Sequence Information of the Target Oligonucleotide and Corresponding probes.

| Name | Sequence (5’-3’) |
| --- | --- |
| Target | CCCAGCGCTTCAGCGTTCTTCGGAATGTCGCGCATT |
| Probe A | ^[a]^AAAAAAAAAAAAAAAAATGCGCGACATTCCGAA |
| Probe B | AAAAAAAAAAAAAAAGAACGCTGAAGCGCTGGG |

[a]: The poly A tail included in the oligonucleotides is used to increase flexibility of the sequence.

**Table S2**. Sequence Information of the Primers, Enzyme Cutting Sites on the RT-LAMP Product, ssDNA as a Target for DNA Hybridization after the Heat Denaturation, and Probes shown in Figure 4A.

| **Name** | **Subtype** | **Sequence (5’-3’)** |
| --- | --- | --- |
| Primers | F3 | GCTGCTGAGGCTTCTAAG |
|  | B3 | GCGTCAATATGCTTATTCAGC |
|  | FIP | GCGGCCAATGTTTGTAATCAGTAGACGTGGTCCAGAACAA |
|  | BIP | TCAGCGTTCTTCGGAATGTCGCTGTGTAGGTCAACCACG |
|  | FLP | CCTTGTCTGATTAGTTCCTGGT |
|  | BLP | TGGCATGGAAGTCACACC |
| dsDNA |  | ^[a]^CCGCAAATTGCACAATTTGCCCCCAGCGCTTCAGCGTTCTTCGGAATGTCGCGCATTGGCATGGAAGTCACACCTTCGGGAACGTGGTT |
| ssDNA |  | CAAATTGCACAATTTGCCCCCAGCGCTTCAGCGTTCTTCGGAATGTCGCGCATTGGCATGGAAGTCACACCTTCGGGAACGTG |
| Probes | A | ^[b]^AAAAAAAAAAAAAAACACGTTCCCGAAGGTGTGACT |
|  | B | AAAAAAAAAAAAAAATCCATGCCAATGCGCGACATT |
|  | C | AAAAAAAAAAAAAAACCGAAGAACGCTGAAGCGCTG |
|  | D | AAAAAAAAAAAAAAAGGGGCAAATTGTGCAATTTG |

[a]: Red letters highlight the region cut by enzymes. [b]: The poly A tail included in the oligonucleotides is used to increase flexibility of the sequence.
